# Gene Sequence to 2D Vector Transformation for Virus Classification

**DOI:** 10.1101/2024.03.12.24304158

**Authors:** Ignacio Sanchez-Gendriz, Karolayne S. Azevedo, Luísa C. de Souza, Matheus G. S. Dalmolin, Marcelo A. C. Fernandes

## Abstract

**Background:** DNA sequences harbor vital information regarding various organisms and viruses. The ability to analyze extensive DNA sequences using methods amenable to conventional computer hardware has proven invaluable, especially in timely response to global pandemics such as COVID-19.

**Objectives:** This study introduces a new representation that encodes DNA sequences in unit vector transitions in a 2D space, extracted from the 2019 repository Novel Coronavirus Resource (2019nCoVR). The main objective is to elucidate the potential of this method to facilitate virus classification using minimal hardware resources. It also aims to demonstrate the feasibility of the technique through dimensionality reduction and the application of machine learning models.

**Methods:** DNA sequences were transformed into two-nucleotide base transitions (referred to as ‘transitions’). Each transition was represented as a corresponding unit vector in 2D space. This coding scheme allowed DNA sequences to be efficiently represented as dynamic transitions. After applying a moving average and resampling, these transitions underwent dimensionality reduction processes such as Principal Component Analysis (PCA). After subsequent processing and dimensionality reduction, conventional machine learning approaches were applied, obtaining as output a multiple classification among six species of viruses belonging to the coronaviridae family, including SARS-CoV-2.

**Results and Discussions:** The implemented method effectively facilitated a careful representation of the sequences, allowing visual differentiation between six types of viruses from the Coronaviridae family through direct plotting. The results obtained by this technique reveal values accuracy, sensitivity, specificity and F1-score equal to or greater than 99%, applied in a stratified cross-validation, used to evaluate the model. The results found produced performance comparable, if not superior, to the computationally intensive methods discussed in the state of the art.

**Conclusions:** The proposed coding method appears as a computationally efficient and promising addition to contemporary DNA sequence coding techniques. Its merits lie in its simplicity, visual interpretability and ease of implementation, making it a potential resource in complementing existing strategies in the field.

## 1 Introduction

The severe acute respiratory syndrome coronavirus 2 (SARS-CoV-2) originated in Wuhan, China, with the first case reported by the World Health Organization (WHO) on December 31, 2019. It rapidly spread worldwide, reaching pandemic proportions that impacted all continents Zhang et al. [2023]. The WHO declared it a Public Health Emergency of International Concern on January 30, 2020, and officially declared it a global pandemic on March 11 of the same year, posing a significant challenge for governments globally and becoming the first modern pandemic of historical proportions. Its impact on public health was overwhelming, straining healthcare systems and resulting in a staggering number of severe cases and deaths globally. By December 2023, there were over 772 million confirmed cases, causing nearly seven million deaths worldwide, as reported by the World Health Organization (WHO) WHO [2023].

Simultaneously, restrictive measures and business closures triggered an unprecedented socioeconomic crisis, leading to massive job losses and a severe impact on the global economy, particularly affecting small businesses and informal workers Vieira et al. [2021], Coccia [2023]. However, the consequences of the pandemic extend beyond health and the economy. The closure of schools and universities disrupted the educational flow, exacerbating learning gaps, especially for those without adequate access to digital resources Kostina and Orlova [2022]. To mitigate the devastating impacts of the pandemic, global leaders and scientists have adopted various measures, including expanded vaccination campaigns and hygiene initiatives. These actions play a crucial role in combating misinformation while strengthening collective immunity and reducing the spread of more virulent variants and their ability to evade immune protection provided by different vaccine types.

Genomic tracking and surveillance of the virus play a crucial role in pandemic management for several reasons. Firstly, these efforts allow health authorities to quickly detect and respond to new outbreaks caused by emerging variants Tosta et al. [2023]. By closely monitoring the virus’s spread and its variants, authorities gain crucial information about transmission patterns, enabling more targeted interventions. Genomic sequencing of SARS-CoV-2 not only aids in identifying new variants but also provides insights into their behavior, transmissibility, and response to COVID-19 treatments. This process plays a fundamental role in tracing virus transmission pathways, allowing specific interventions such as the development and adjustments of diagnostic tests, both serological and molecular, as well as potential updates in vaccine formulations and early detection of variants of concern (VOCs), variants of interest (VOIs), and variants under monitoring (VUM) Tosta et al. [2023].

Global collective efforts in identifying and mitigating the disruptions caused by SARS-CoV-2 have led to the creation of extensive open repositories of viral genomic sequences, shared globally. This has enabled the generation of an unprecedented number of genomic sequences, facilitating the classification and study of this virus almost in real-time. Viral classification provides a systematic framework supporting multidisciplinary research in epidemiology, virology, and public health, contributing to the understanding of the virus’s characteristics and the implementation of specific measures. Additionally, comprehensive tracking of these variants, fueled by a robust taxonomic classification, provides valuable data for predictive modeling, establishing itself as an indispensable resource in anticipating and addressing future pandemics Levi et al. [2024].

While sequencing SARS-CoV-2 is crucial for understanding the virus and its variants, it faces a series of challenges Chiara et al. [2021], De Maio et al. [2020]. The substantial volume of data from sequencing can overwhelm existing computational and analytical capabilities, especially when dealing with the highly mutable RNA virus, consisting of approximately 30,000 base pairs. In bioinformatics, the practice of assigning a genomic sequence to a given group is mostly based on similarities in the structure and organization of the genomes of the studied species, and this classification contributes to their phylogenetic and functional understanding. Tasks such as alignment, assembly, and comparison of long sequences require significant computational power Phan et al. [2015]. As the sequence length increases, traditional algorithms may become computationally intensive and less effective Sf [1990], Hofstetter et al. [2019]. The National Center for Biotechnology Information (NCBI) host a sequence alignment-based approach, BLAST Sf [1990], which searches similarity in samples. This technique has been the main tool for genomics research in bioinformatics for the last 30 years Shah et al. [2019]. However, BLAST exhibits certain limitations, as noted in the study Hofstetter et al. [2019], which employed this tool to identify fungi collected in a protected beech forest in Montricher, Switzerland. The research highlights that the accurate interpretation of results obtained through BLAST alignments demands extensive taxonomic and molecular expertise. Additionally, it emphasizes that such analyses are excessively time-consuming for routine application in future applications, exemplified by the two-month duration required for verifying approximately 20 identifications.

Given the vast amount of genomic data available, scientists have leveraged the power of machine learning (ML) and deep learning (DL) algorithms in various aspects, including sequence classification and pandemic monitoring, to assist in combating the virus’s spread Sahoo et al. [2023]. However, the complexity and variability of genomic data pose significant challenges in extracting and interpreting features, as well as in the high computational cost required for manipulation, often hindering the viable use of traditional machine learning algorithms Naeem et al. [2021], Singh et al. [2021a]. The way genomic data is preprocessed and transformed before being fed into machine learning techniques influences the classification results. The ViraMiner Tampuu et al. [2019] project is an example of the application of sequences in text form, meaning that the proposed methodology uses raw DNA data to identify potential viral sequences across human samples. The classification model employs two branches of convolutional neural networks (CNN) to detect patterns and frequency patterns on metagenomics contigs, achieving a 0.923 area under the receiver operating characteristic (ROC) curve, and precision of 90%. However, he work presented in Habib et al. [2020] introduced the COVIDier software for virus classification. Instead of utilizing raw sequences, they employed a CounterVectorizer function to convert the 1925 genome sequences of six species in the Coronavirus family into numeric information. The machine learning algorithm used was the Multi-layer Perceptron Classifier, which achieved a precision of 99%.

Therefore, the quality of genomic data representation is crucial to obtain precise and meaningful insights through these models de Souza et al. [2023], Naeem et al. [2021]. Various techniques are employed to represent this data before applying these models, considering that nucleotides in sequences, represented by characters (usually A, T, G, and C), must be numerically represented for subsequent processing de Souza et al. [2023]. Specific techniques in genomic signal processing (GSP), a field of bioinformatics that utilizes algorithms from Digital Signal Processing and sequence analysis, provide methods for representing, analyzing, and interpreting patterns in genetic data Mendizabal-Ruiz et al. [2018], Adetiba et al. [2022]. These techniques find application in various bioinformatics tasks, including viral classification de Souza et al. [2023], exon and intron sequence classification Abo-Zahhad et al. [2012], Bonidia et al. [2021a], identification of biomarkers Kung et al. [2010], and identification of protein-coding regions. Considering this, Naeem et al. [2021] applied the GSP techniques discrete Fourier transform (DFT), discrete cosine transform (DCT), and seven moment invariants was employed for feature extraction from three species of human strains of the Coronavirus. Two classifiers were utilized, namely KNN and the trainable cascade-forward backpropagation neural network. The KNN model achieved accuracy and F1 score of 100%, training with only 46 complete genomes for each species. In the study presented in Hoang et al. [2016], the authors employed Frequency Chaos Game Representation to convert genome sequences into images. Additionally, they utilized DCT and Singular Values Decomposition to reduce the dimensionality of the images into vectors. These vectors were then used for rapid sequence retrieval, the construction of phylogenetic trees, and the classification of viral genomic data.

In the context of using GSP for the analysis and feature extraction of genomic sequences, the application of the Fourier Transform to convert sequences from the time domain to the frequency domain stands out as one of the most popular and powerful tools within GSP. The work described in Hoang et al. [2016] utilized Chaos Game Representation (CGR) for analyzing the DNA sequences of various human-infecting viruses. It then applied the DFT to these sequences and calculated the corresponding power spectra for each. To standardize the sequence lengths, an even scaling method was applied across all sequences. The primary aim of this research was to construct phylogenetic trees to classify genes and genomes of virus subtypes.

Applying machine learning techniques associated with GSP for viral classification, the work Randhawa et al. [2020] mapped genomic sequences of the Riboviria realm virus into two-dimensional data using the CGR. Subsequently, it obtained the magnitude spectrum of the signal through the DFT and constructed a pairwise distance matrix with the Pearson Correlation coefficient. The ML algorithms used in this application were Linear Discriminant, Linear and Quadratic SVM, Fine KNN, and Subspace Discriminant and Subspace KNN, with a 10-fold cross-validation. The best model was Quadratic SVM, which achieved an accuracy of 94.9% in identifying the family and realm of the SARS-CoV-2 virus. The work in de Souza et al. [2023] also applied this two concepts into their research, using CGR, magnitude and phase spectrum of the DFT to propose a genome representation to map SARS-CoV-2 and other five species from Coronaviredae family, obtaining three reduced signatures. Used a unidimensional CNN as classifier, achieving a accuracy of 99.69%.

This manuscript introduces an innovative representation method that encodes viral DNA sequences into unit vector transitions within a two-dimensional space. Engineered to optimize virus classification workflows, this strategy ensures efficient and precise virus identification with minimal demands on hardware resources. Leveraging GSP techniques, the manuscript unveils a novel approach for converting DNA sequences into two-dimensional numerical representations. It aims to validate the efficacy of traditional machine learning models in accurately differentiating among six virus types from the Coronaviridae family, including SARS-CoV-2. Characterized by its straightforwardness, visual clarity, and ease of deployment, the proposed method is poised to refine existing viral classification frameworks and genetic sequence analyses. Addressing the urgent need for rapid responses to global health crises like COVID-19, this research contributes a computationally lean solution for the in-depth examination of DNA sequences, offering valuable insights for practical deployment in various real-world contexts.

The main contributions of this study are outlined as follows:

- **Introduction of a Novel DNA Encoding Method**: We introduce a pioneering approach that encodes viral DNA sequences into unit vector transitions within a two-dimensional space, thereby enhancing the efficiency and interpretability of genetic data analysis.
- **Enhanced Virus Classification**: The method’s efficacy in accurately distinguishing between six virus types within the Coronaviridae family, including SARS-CoV-2, is demonstrated and supported by high-performance metrics.
- **Computational Efficiency**: The study achieves notable computational efficiency by requiring minimal computational resources for comprehensive genetic analyses and virus classification, making it particularly valuable in resource-constrained settings and aiding in global viral outbreak surveillance and management.
- **Simplicity and Accessibility**: The method is characterized by its simplicity, interpretability, and ease of implementation, making it accessible to a broad audience, including those without specialized knowledge in GSP, and facilitating its adoption in various research and practical applications.
- **Implications for Public Health and Bioinformatics**: The contributions of this study are poised to significantly influence both bioinformatics research and public health strategies for pandemic response. By providing an efficient and precise tool for viral classification and DNA sequence analysis, the work enhances the capacity for rapid viral identification and outbreak monitoring.

The structure of the article is outlined as follows: Section II provides a comprehensive description of the methodologies and techniques employed in this research. In Section III, the findings are presented and discussed, including a comparative analysis with existing state-of-the-art results. The article concludes with final remarks in Section IV.

## 2 Materials and Methods

This section delineates the methodology employed for encoding DNA sequences into a two-dimensional space, utilizing twiddle factors obtained from the DFT representation. The procedural workflow is illustrated in Figure 1, providing a succinct and clear overview.

**Figure 1:**
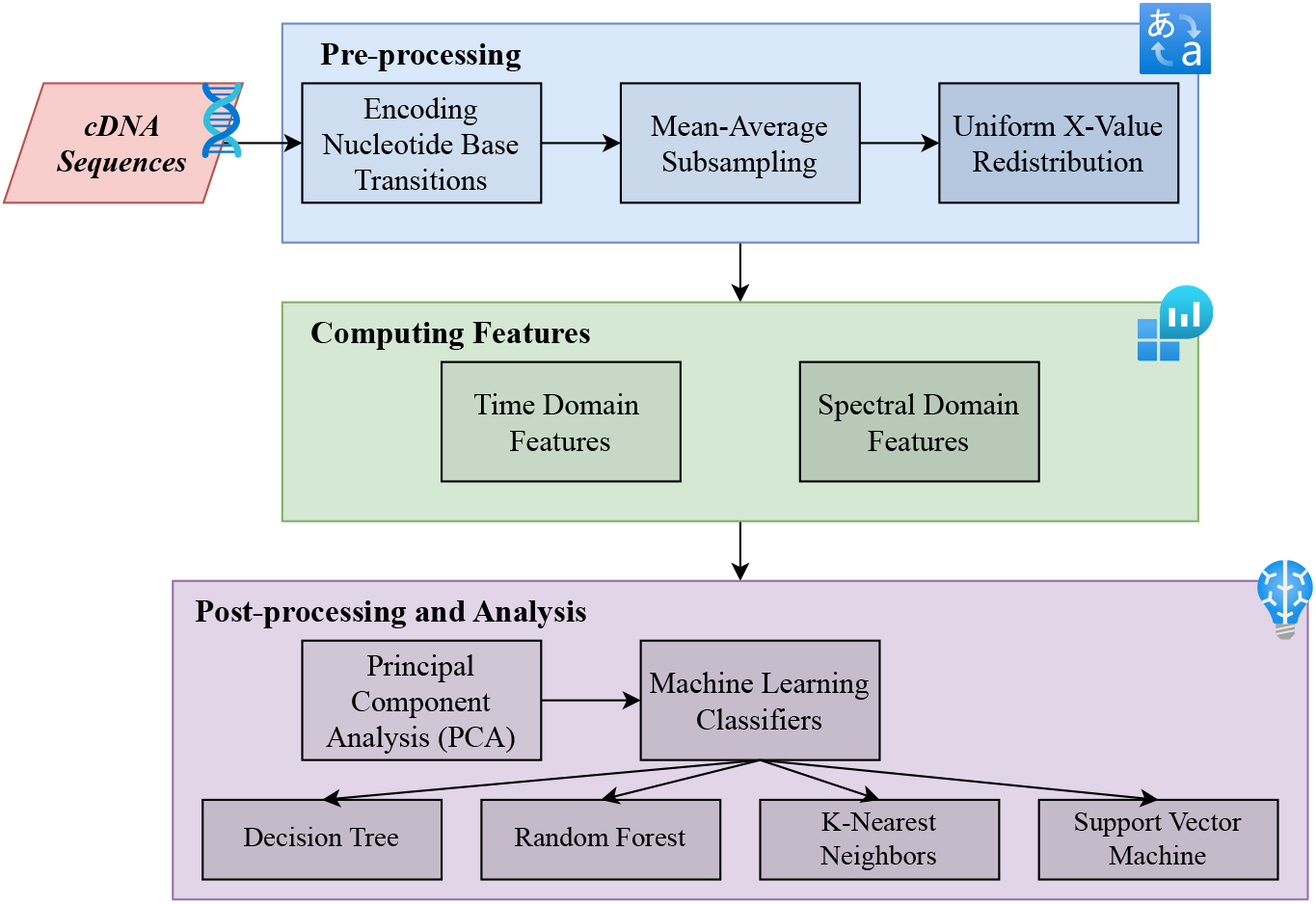
Workflow of the proposed methodology for Gene sequence 2D transformation for virus classification.

### 2.1 Discrete Fourier Transform (DFT) Fundamentals and Twiddle Factors

The DFT is a mathematical tool extensively employed in signal processing to convert one-dimensional numerical sequences into their frequency domain counterparts. The DFT is mathematically expressed as shown in Equation 1:

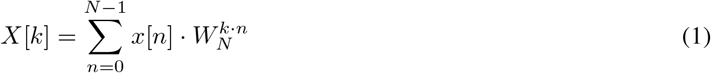

In this equation, *X*[*k*] denotes the *k*-th complex frequency component of the DFT output, and *x*[*n*] represents the *n*-th element of the input data sequence. The term *N* signifies the total number of data points in the sequence. Both indices *k* and *n* range from 0 to *N−*1, facilitating the enumeration of frequency components and data sequence elements, respectively.

The DFT formula presented in Equation 1 is a symplyfied version, which use the concept of the twiddle factor *W*_*N*_ . The term twiddle factor is defined as in Equation 2:

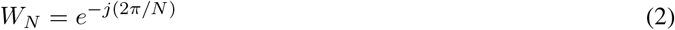

In this representation, *j* is the imaginary unit (*j*^2^ = *−*1), and *e* is the base of the natural logarithm.The term 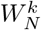 represents the twiddle factor raised to the power of *k*. For a specific *k* value ranging from 0 to 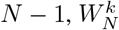 corresponds to a complex number on the unit circle in the complex plane.

The magnitude 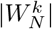 is always equal to 1, indicating that it represents a unit vector on the complex plane. The phase component *e*^*jθ*^ determines the angle *θ*, which positions the complex number on the unit circle.

As *k* varies from 0 to *N−*1, the angle *θ* changes, causing the complex number represented by 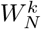 to rotate on the unit circle. For example, if *N* = 16 and *k* varies from 0 to 15, 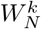 represents complex numbers on the unit circle with different angular positions. The phase (angular values) of the 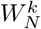 vectors can be determined as *θ* = 2*πk/N* .

### 2.2 Representing Nucleotide Transitions with Twiddle Factors

In this study, we assert that transitions between nucleotide bases can be effectively represented by corresponding 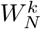 complex values, which are unit vectors. This assertion constitutes a significant and foundational aspect of our encoding method. For an illustration of the 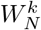 vectors, refer to Figure 2. The methodology for encoding DNA sequences into a two-dimensional numerical space is summarized as follows:

**Figure 2:**
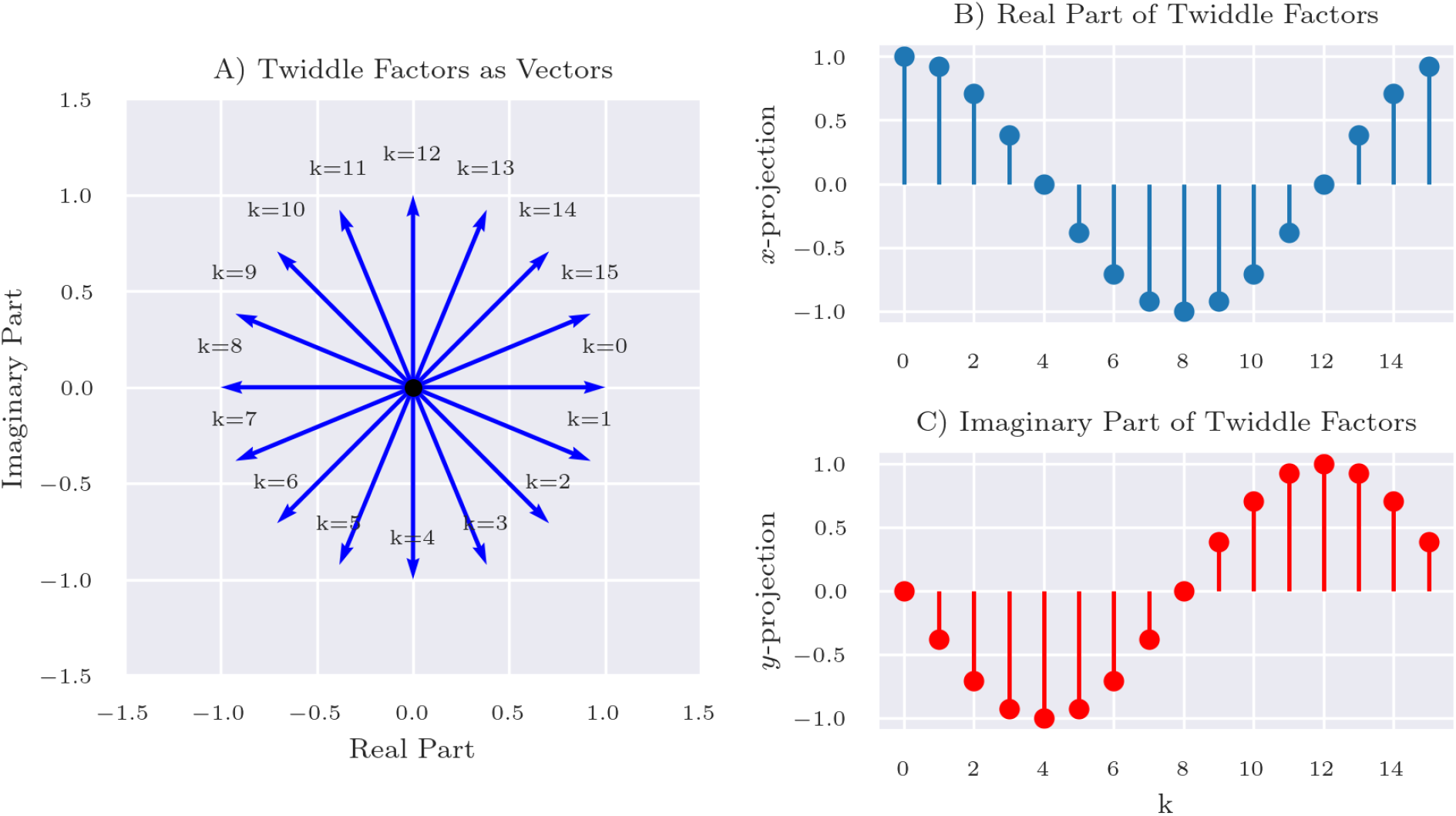
Visualization of Twiddle Factors used in encoding transitions between nucleotide bases. Panel A visually depicts the twiddle factors as vectors in the complex plane, annotated with their respective *k* values. Panels B and C exhibit the real and imaginary components of these twiddle factors, respectively.

- **Sequence Transition**: The DNA sequence of viruses, comprising a series of nucleotides, is initially translated into a sequence of transitions. Each transition encapsulates a pair of consecutive nucleotides, capturing the sequential information inherent in the DNA sequence.
- **Mapping Transitions to Indices**: Employing a predefined encoding scheme detailed in Table 1, each unique nucleotide transition is allocated a distinct numerical index. This step converts the sequence of nucleotide pairs into an index sequence, setting the stage for subsequent encoding steps.

**Table 1:** Transitions between Nucleotide Bases
- **Applying Twiddle Factors**: Twiddle factors, are then assigned to each numeric transition based on the index derived from the encoding scheme. Figure 2 graphically represents these associations. These twiddle factors are instrumental in transposing transitions into a two-dimensional numerical space.
- **Encoding into 2D Space**: Commencing from the origin (0,0) in the 2D space, the encoding process iteratively applies the twiddle factor corresponding to each transition to navigate to the subsequent point. This recursive addition of twiddle factors allows for the construction of a path through the 2D space, ultimately encoding the entire sequence of transitions as a path in the 2D plane.

|  | A | C | G | T |
| --- | --- | --- | --- | --- |
| A | $A \rightarrow A^0$ | $A \rightarrow C^4$ | $A \rightarrow G^8$ | $A \rightarrow T^{12}$ |
| C | $C \rightarrow A^5$ | $C \rightarrow C^1$ | $C \rightarrow G^9$ | $C \rightarrow T^{13}$ |
| G | $G \rightarrow A^6$ | $G \rightarrow C^{10}$ | $G \rightarrow G^2$ | $G \rightarrow T^{14}$ |
| T | $T \rightarrow A^7$ | $T \rightarrow C^{11}$ | $T \rightarrow G^{15}$ | $T \rightarrow T^3$ |

This representation is central to our methodology for encoding DNA sequences. Figure 3 outlines the novel encoding technique, which forms the essence of our innovative approach. Panel A of Figure 3 depicts a complete DNA sequence of a virus in two-dimensional space, illustrating the dynamic representation of the sequence and offering insight into its underlying patterns and nucleotide transitions. Panel B presents a specific segment of the DNA sequence, detailing the sequence of base transitions alongside their corresponding numerical indices, thereby shedding light on the foundational mapping and conversion processes integral to our encoding strategy. Finally, Panel C demonstrates the conversion of a DNA sequence segment into vectors within the two-dimensional plane, emphasizing the transformative aspect of our approach which simplifies the visualization and analysis of complex genetic information.

**Figure 3:**
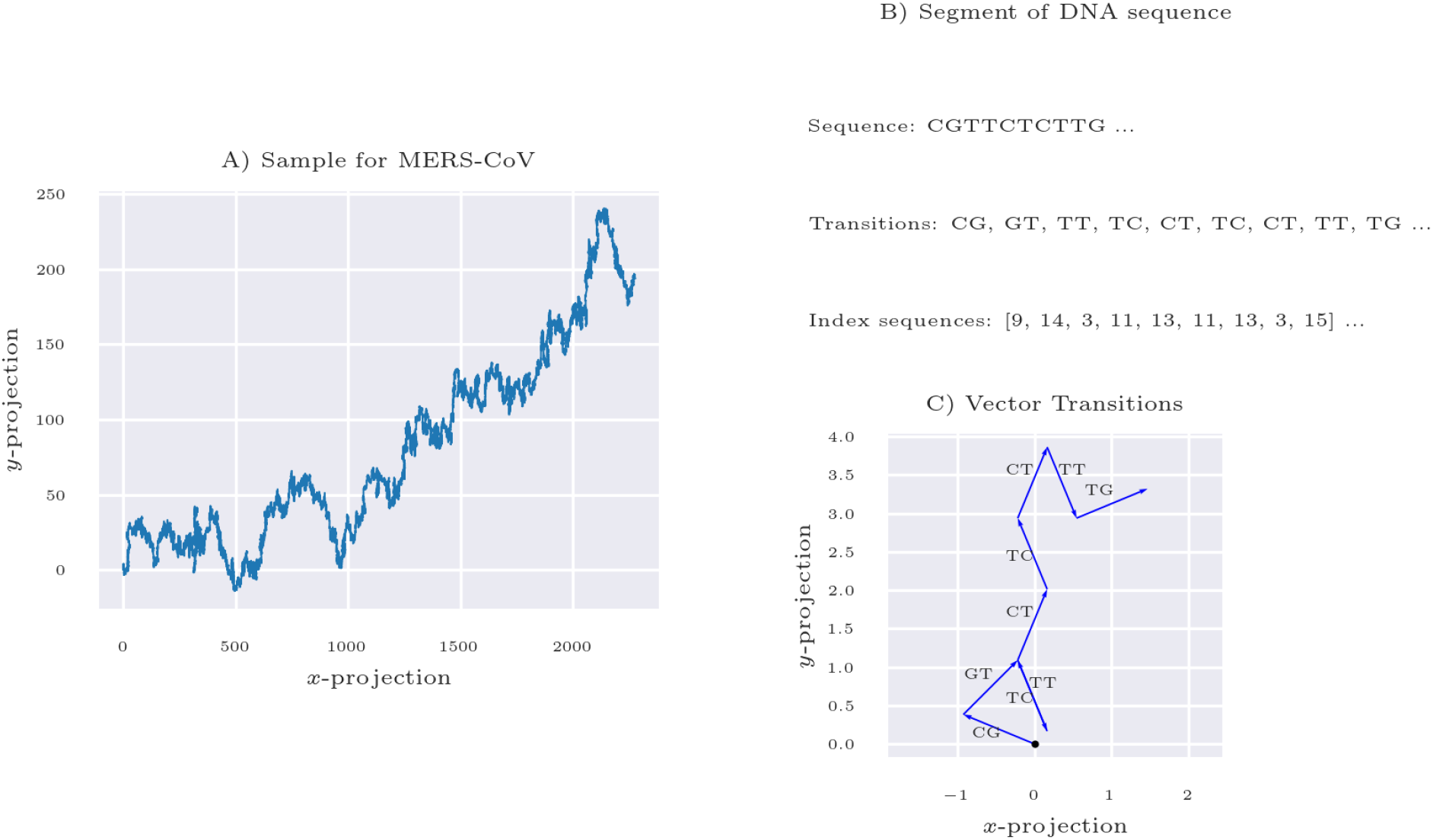
Transformation of DNA sequences into 2D vector transitions. Panel A presents a complete DNA sequence of a virus, dynamically unfolded in a two-dimensional space, revealing intricate patterns and relational nuances inherent in the sequence. Panel B focuses on a segment of the DNA sequence, disclosing individual base transitions and their corresponding sequential indices, elucidating the core mechanics of our encoding strategy. Panel C showcases the conversion of a DNA sequence segment into expressive vectors in the two-dimensional domain, highlighting the innovative transformation process that simplifies the visualization and analysis of complex genetic data.

In subsequent sections, we will harness this encoding method in conjunction with subsequent processing, further dimensionality reduction and Machine Learning techniques to explore its effectiveness in distinguishing between six viral species within the Coronaviridae family.

### 2.3 Pre-processing

#### 2.3.1 Encoding Nucleotides Base Transitions

In molecular biology, nucleotide bases are commonly represented as character vectors, with each letter corresponding to adenine (A), cytosine (C), guanine (G), or thymine (T). Consequently, there are only 16 possible transitions between these bases, as outlined in Table 1. It is essential to highlight that encoding specific nucleotide base transitions to particular indexes *k* = 0, 1, 2 … 15 encompasses numerous possible combinations. In this study, a selective combination is employed with the aim that transitions from one base to another should not use consecutive indexes; for instance, *C→A* is assigned an index of 5, *C→C* an index of 1, *C→G* an index of 9, and *C→T* an index of 13. However, although exploring all possible combinations is beyond the scope of this study, alternative strategies for encoding base transitions to indexes remain a promising area for future research.

#### 2.3.2 Mean-average subsampling

After transforming the DNA sequences into numerical points in 2D space, a subsequent processing step known as “mean-average subsampling” is applied to enhance the data representation. This step aims to encapsulate the broader trends of the sequences in the 2D numerical space, allowing minor details, such as backward transitions, to be deprioritized. In this context, the sequences in the 2D space are organized in ascending order based on their x-values (abscissa), ensuring a coherent progression in the data representation.

In the mean-average subsampling process, a window encompassing *M* points is utilized. The operation commences from the second point in the sequence, keeping in mind that each frequency originates from the point (0,0). The window moves across the sequence, and within each window, the mean average of the points is computed, yielding a single point representative of that window. It’s crucial to note that the window moves without overlap; each subsequent window begins after the end of the previous window, ensuring unique sets of points for each calculation. This step repeats iteratively, traversing through the sequence until all points are processed.

The result of the subsampling process is a collection of new numerical sequences that have been derived through subsampling from the original data. These refined sequences maintain the fundamental attributes of the initial data, yet they exhibit a dimensionality reduction. This streamlined dimensionality facilitates enhanced simplicity and efficiency in subsequent analyses, as well as more coherent visual representations in the 2D space.

#### 2.3.3 Uniform X-Value Redistribution

In the data preprocessing phase, a uniform x-value redistribution technique was applied to the 2D numerical sequences. This method aimed to equalize the intervals of x-values (abscissae) across the entire sequence, ensuring a more systematic and uniform distribution. Each x-value was recalculated based on a mean interval value, resulting in a sequence that spans uniformly from the initial to the final original x-value.

The y-values (ordinates) of the sequence remained unaltered during this process, maintaining the integrity of the original data. This step ensures that the redistributed sequences are more analyzable, offering a consistent basis for subsequent data interpretation and analysis in the 2D space.

### 2.4 Computing Features from ‘Time’ and ‘Spectral Domain’

Dimensionality reduction strategies, which encompass both feature selection and feature extraction, have been advanced to address the complexities inherent in genomic data Afshar and Usefi [2020], Bonidia et al. [2021b]. These strategies are pivotal when working with high-dimensional datasets, such as those derived from genomic sequencing.

In this context, features for classifying virus sequences were computed using the two-dimensional space representations of preprocessed sequences. These representations were obtained subsequent to the application of mean-average subsampling (Section 2.3.2) and Uniform X-Value Redistribution (Section 2.3.3). The primary objective of this computation is not merely to optimize feature extraction but to underscore the effectiveness of our DNA sequence encoding methodology. Our focus is to derive features that unequivocally illustrate the capacity of our encoding strategy for efficient low-dimensional representation, facilitating tasks such as virus classification and visual discrimination.

Two distinct categories of features were extracted for this purpose: time-domain features and spectral-domain features. Each category is elucidated in the following sections, providing insights into their computation and relevance to the overarching goal of the study.

#### 2.4.1 Time Domain Features

Time domain features are derived directly from the preprocessed sequences in the 2D space. Observing the curves, we can see clear differences in the transitions between various viruses. To get these features, a second-degree polynomial fit and Linear Predictive Coding (LPC) Spratling [2017] with three coefficients were applied. LPC, often used in speech processing, helped in obtaining useful features directly from the sequence curves. These features are organized into a matrix that will be used later for dimensionality reduction and classification.

#### 2.4.2 Spectral Domain Features

Spectral domain features were also extracted to complement the time domain features, the DFT was employed for this purpose. Before applying DFT, the sequences were adjusted to meet its assumptions. Specifically, the y-values from the sequences in the 2D space were manipulated to represent a period of a supposed periodic sequence. This adjustment involved flipping the sequence and concatenating it with the original, preparing the sequences adequately for DFT application. Upon the application of DFT, we retained the magnitudes of the coefficients at specific positions, choosing indices one to three, while the zero index was disregarded because it contains merely the mean value information.

Following the extraction of spectral and time domain features, a comprehensive representation is created, which streamlines the subsequent stages of analysis and classification. Post feature extraction, Principal Component Analysis (PCA) is employed to reduce the dimensionality of the feature space Howley et al. [2006], mapping each sequence to a unique point in a new, simplified space. This refined representation facilitates enhanced visualization and the application of conventional Machine Learning (ML) techniques for more effective classification in subsequent analyses.

### 2.5 Experimental Setup

In this section, we elucidate the methodology adopted in our experiments, describing the datasets, machine learning models, evaluation metrics, and validation strategies that constitute the backbone of our research.

#### 2.5.1 Datasets

Our experiments leverage datasets composed of gene sequences meticulously sourced from various viruses. It contains 12,467 viral sequences from six species of the Coronaviridae family, namely Severe Acute Respiratory Syndrome-related Coronavirus (SARS-CoV-2), Betacoronavirus 1, Middle East Respiratory Syndrome-related Coronavirus (MERS-CoV), Human Coronavirus NL63 (HCoV NL63), Human Coronavirus 229E (HCoV 229E), and Human Coronavirus HKU1 (HCoV HKU1). Encompassing a spectrum of virus types, each contributing to a comprehensive repository, which is instrumental for a robust and extensive analysis. These samples were obtained from the National Genomics Data Center (NGDC), and contain sequences from 67 countries with a genome length ranging from 26000 to 32000 bases par (bp). A filter was applied to select only complete samples with an N’s number less than 0.01%. This carefully curated collection of gene sequences is pivotal in ensuring the accuracy and reliability of our experimental outcomes.

#### 2.5.2 Machine Learning Models

In our study, we employ a selection of machine learning models to classify virus sequences, each with its unique foundational principles and computational mechanics.

##### Decision Tree (DT)

A Decision Tree is a supervised learning algorithm widely used in data mining due to its simplicity and ease of interpretation Loyola-González et al. [2023]. This algorithm operates on a set of logical rules, requiring minimal parameter tuning to process diverse data types effectively Su and Zhang [2006]. The construction of a Decision Tree begins with selecting an initial criterion, such as entropy reduction or information gain, to determine the best splitting attribute for the root node Somvanshi et al. [2016]. Based on this attribute, the training data is segmented into subsets, each corresponding to a branch leading to a decision node. The algorithm continues this attribute selection process, data segmentation, and node splitting, progressively expanding the tree with new branches and nodes. This iterative process culminates in forming leaf nodes, each representing a class label—effectively the algorithm’s decision output Su and Zhang [2006], De Ville [2013]. By iteratively partitioning the data in this manner, the Decision Tree algorithm efficiently classifies the input data, making it a powerful tool for predictive modeling.

##### Random Forest (RF)

An Ensemble of Decision Trees - stands as a robust ensemble method, particularly adept for both regression and classification tasks, including scenarios with multiple classes. It’s renowned for its straightforward and swift deployment, making it a favored choice in handling datasets characterized by high dimensionality and substantial volume Breiman [2001]. Within the realm of classification, the algorithm sets its sights on crafting a predictive function *f* (*X*), with *X* representing the input vector and *Y* denoting the anticipated output. The essence of the algorithm lies in minimizing the discrepancy between *Y* and *f* (*X*), quantified by the error function *L*(*Y, f* (*X*)). This is achieved by amalgamating a suite of base learners, essentially individual decision trees *h*_1_(*x*), …, *h*_*j*_(*x*), that collectively cast their votes to predict the most likely class for a given input *x ∈ X* Cutler et al. [2012]. *The predictive function f* (*X*) for classification is thus formulated as:

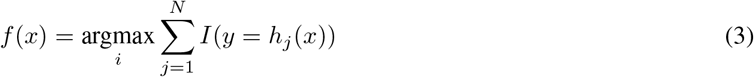

Here, argmax identifies the class index *i* that maximizes the sum, *N* signifies the total number of classes, and *I* is the indicator function, which assigns a value of 1 when the condition *y* = *h*_*j*_(*x*) holds true for *y ∈ Y*, and 0 otherwise. This collaborative voting mechanism of the decision trees enhances the algorithm’s accuracy and reliability in predictions, showcasing the power of ensemble learning.

##### K-Nearest Neighbors (KNN)

KNN is a non-parametric, instance-based learning algorithm. It classifies a data point based on how its neighbors are classified, utilizing distance metrics, such as the Euclidean distance, to obtain dissimilarities between examples in a given feature space. The class to which a test example *r*_*i*_ belongs will be determined through a majority vote among the K nearest neighbors, as seen in:

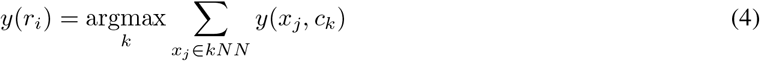

where *x*_*j*_ is a *k* nearest neighbor of the training set, and *y*(*x*_*j*_, *c*_*k*_) indicates if *x*_*j*_ belongs to the *c*_*k*_ class Sun and Huang [2010]. KNN is inherently adaptive, capable of updating its classification decision as the dataset evolves, providing a level of flexibility that is essential when dealing with dynamic gene sequence data.

##### Support Vector Machine (SVM)

SVM is a powerful model known for its effectiveness in classification tasks. It operates by finding the hyperplane that distinctly classifies data points in a multi-dimensional space, ensuring the maximization of the margin between classes. The equation for the optimal hyperplane is given by:

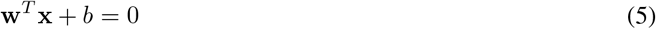

where **w** is the adjustable weight vector, **x** is the input vector, and *b* is a bias, where the algorithm has to find **w** and *b* that will make the hyperplane best separate the data. The algorith SVM is renowned for its capacity to manage high-dimensional data, making it especially pertinent for the classification of intricate gene sequences Huang et al. [2018].

#### 2.5.3 Evaluation Metrics

Our experiment’s integrity and the reliability of results are fortified by employing a comprehensive set of evaluation metrics such as accuracy (Equation 6), precision (Equation 7), recall (Equation 8), and F1-score (Equation 9) Sokolova and Lapalme [2009]. These metrics furnish a multi-dimensional perspective, allowing for a meticulous assessment of the models’ performance in classifying virus sequences.

Accuracy quantifies the overall correctness of a model across all classes, offering a broad perspective on the model’s efficacy based on the ratio of correct predictions to the total number of predictions. It encapsulates the model’s precision in generating optimistic predictions and its proficiency in minimizing false positives. On the other hand, Recall emphasizes the model’s success in identifying all true positives, thereby underscoring its effectiveness in reducing false negatives. The F1-score, calculated as the harmonic mean of precision and Recall, balances these two metrics. It emerges as a pivotal metric when there is an imperative to harmonize sensitivity with precision. Specifically, in the context of viral classification, a high recall value indicates the model’s adeptness at accurately identifying actual positive cases, crucially mitigating the incidence of false negatives. Such accuracy is paramount in curbing the dissemination of diseases by enhancing the detection of actual viral infections. The Accuracy, Precision, Recall, and F1-score can be expressed as:

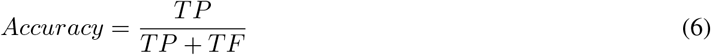

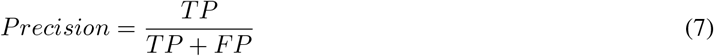

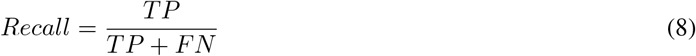

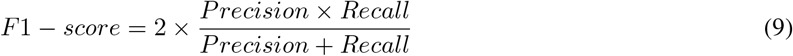

where TP corresponds to the number of true positive samples, FP to the number of false-positive samples, FN number of false negative specimens and TN number of true negative specimens.

#### 2.5.4 Validation Strategy

For the training phase, the dataset was segregated into training and validation subsets following an 80% to 20% ratio, with a keen focus on maintaining class stratification. This stratification is crucial, especially for datasets that exhibit class imbalances. Stratified cross-validation was the chosen method, with a ten-fold (*k* = 10) approach determined to be most effective after evaluating various fold configurations for optimal model performance.

Regarding hyperparameters, extensive tuning was considered unnecessary, as the default settings offered by the scikit-learn library were suitable for most models. The settings for the model hyperparameters are outlined as follows:

- **Random Forest (RF):** Settled on using 10 trees in the ensemble, with a requirement of at least 2 samples to split an internal node. This configuration aims to balance model complexity with generalization capability.
- **K-Nearest Neighbors (KNN):** Opted for *k* = 3, meaning the classification of a sample is influenced by the three nearest neighbors, employing the Euclidean distance to measure closeness.
- **Decision Tree (DT):** Imposed a maximum depth of 5 levels for the tree and required a minimum of 5 samples per node to curb the model’s complexity, enhancing its generalization to new data.
- **Support Vector Classifier (SVC):** Adopted a radial basis function (RBF) kernel, with the gamma parameter set to ‘auto’, facilitating an adaptable decision boundary that adjusts to the data’s intrinsic distribution without manual tuning.

These model-specific configurations were strategically chosen to optimize performance while ensuring the models remain robust and generalizable to unseen data. Learning curves for each model were generated to observe their behaviors during the training process as the number of samples increased, as shown in Figure 4.

**Figure 4:**
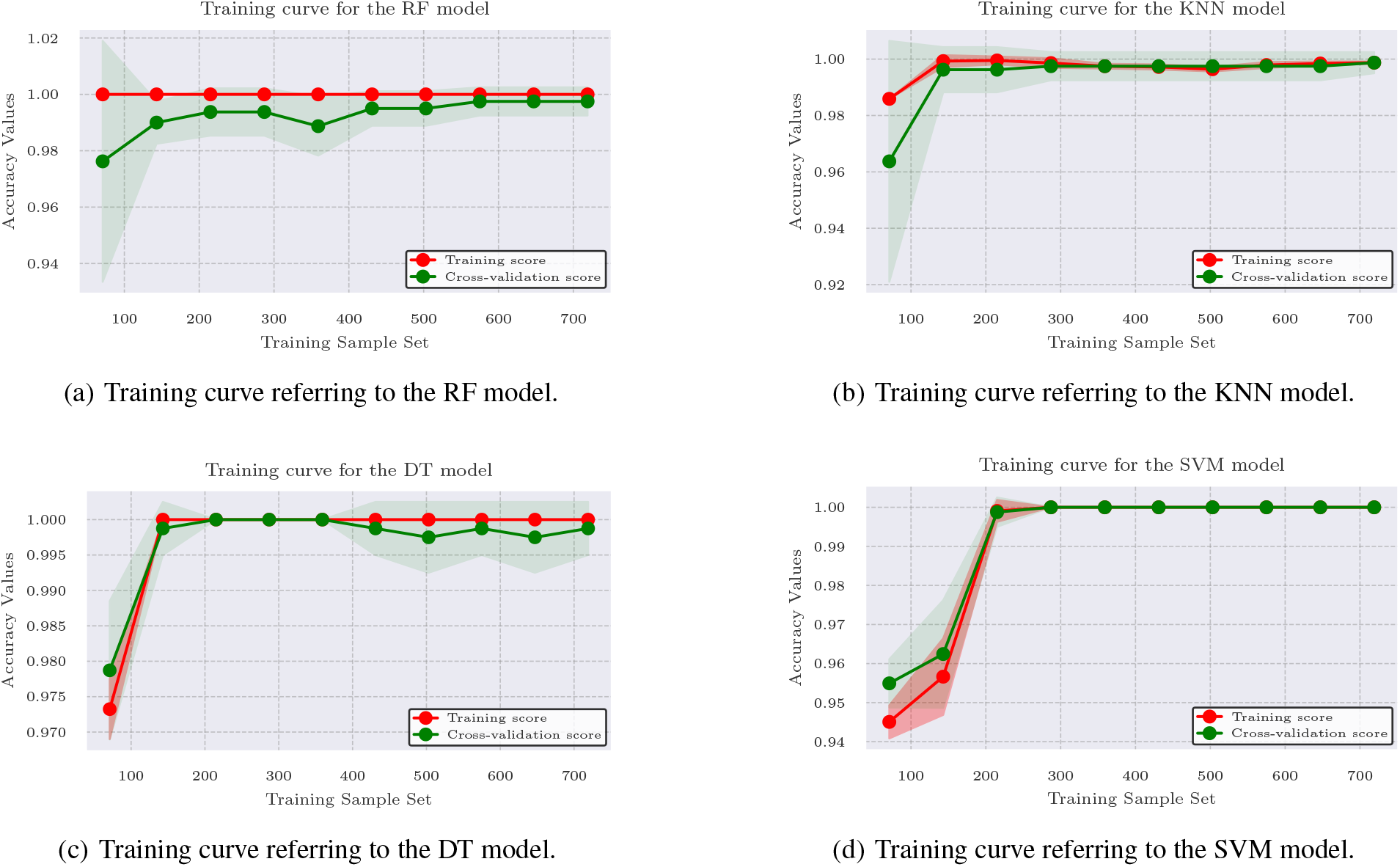
Training curves referring to the ML models adopted in this paper.

It is observed that the training and validation scores remain high (above 90%) in all models. With each iteration, validation scores tend to improve as more training data is incorporated with the exception of the 4(a) curve where there is a slight drop in its score as the amount of interaction and samples increases, however, the validation curve quickly approaches the training curve. The training curve obtained through the DT model (see Figure 4(c)), presents a subtle fluctuation between its validation curve even with the increase in the number of samples, which may indicate potential saturation of the model. All curves indicate that there is a good compensation between bias and variance and indicate that the quantities of samples used in model training would be sufficient, considering that the models stabilize with a considerable number of samples. The results of the analysis of the learning curves indicate a robust and promising performance of the evaluated models. The consistency of high scores in both training and validation suggests an effective learning and generalization capability. The identification of potential saturation points in certain models, particularly notable in the training curve of the DT model, indicates the need to consider additional strategies to optimize performance as the complexity of the problem increases or with the expansion of data. The absence of signs of overfitting and the minimal difference between the training and validation curves reinforce the reliability of the models to accurately generalize to new datasets.

## 3. Results and Discussion

Figure 5 presents the two-dimensional encodings of DNA sequence samples from five distinct viruses. Each panel within the figure corresponds to a different virus type and illustrates a subset of the randomly chosen samples to facilitate comparative analysis. The patterns of transitions in the sequences exhibit notable inter-class variations and consistent intra-class similarities. This observation aligns with the expectation, underscoring the method’s capability to effectively capture and represent the intrinsic information of various DNA sequences associated with different virus types.

**Figure 5:**
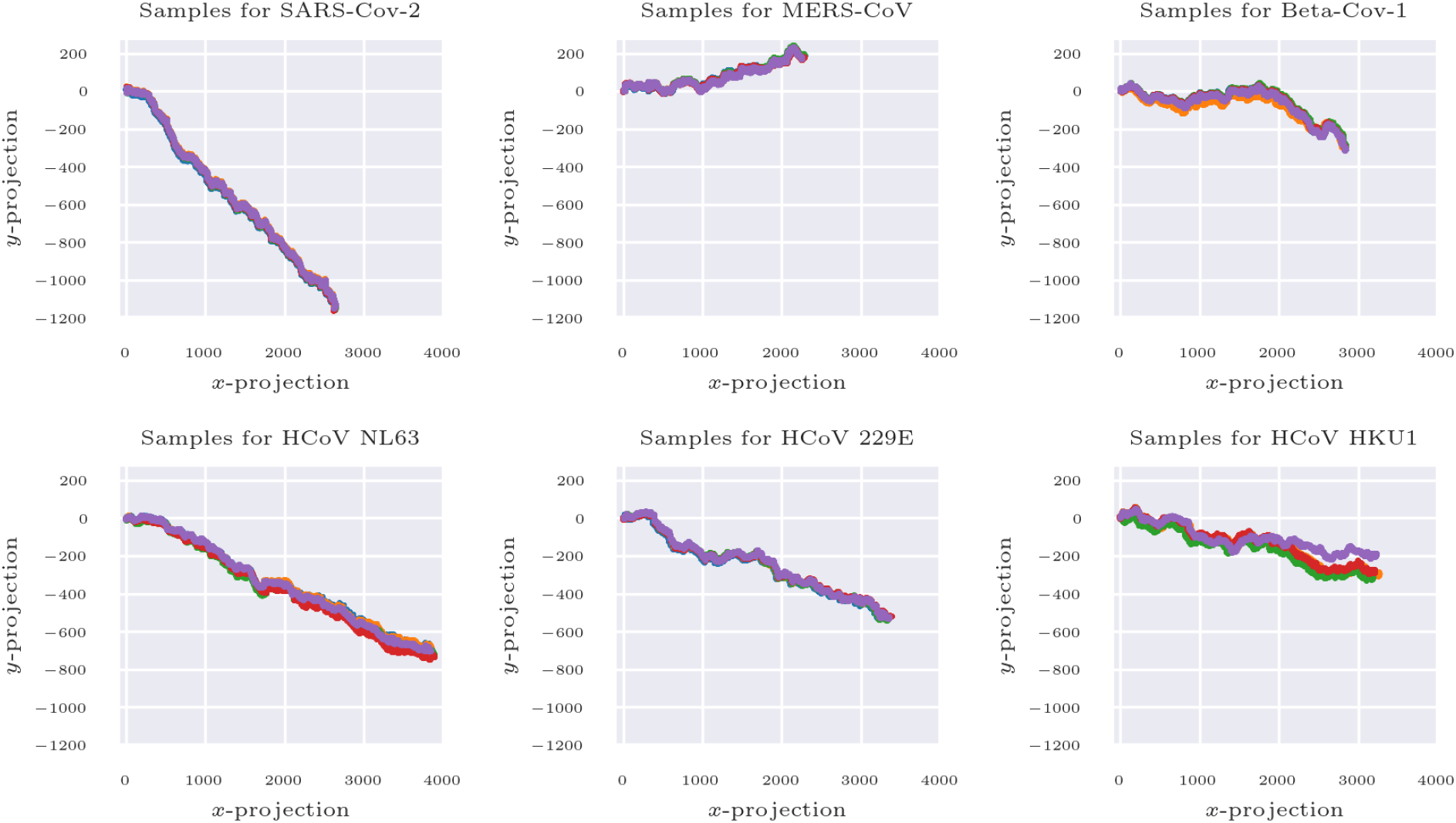
Illustration of encoded DNA sequences from six different virus types. Each panel represents a distinct virus type, displaying five randomly chosen samples for enhanced visual comparison.

Intrigued by the distinctive 2D spatial characteristics, we explored temporal feature extraction as depicted in Figure 6. Polynomial fitting (degree *N* = 2) and Linear Predictive Coding (LPC) were employed to ascertain features commonly associated with the time domain. This initiative aimed to gauge the method’s viability and effectiveness as a precursor to further dimensionality reduction procedures. The sequences were also subject to Discrete Fourier Transform (DFT), retaining coefficients ranging from 1 to 3, following the preprocessing steps outlined in section 2.3.

**Figure 6:**
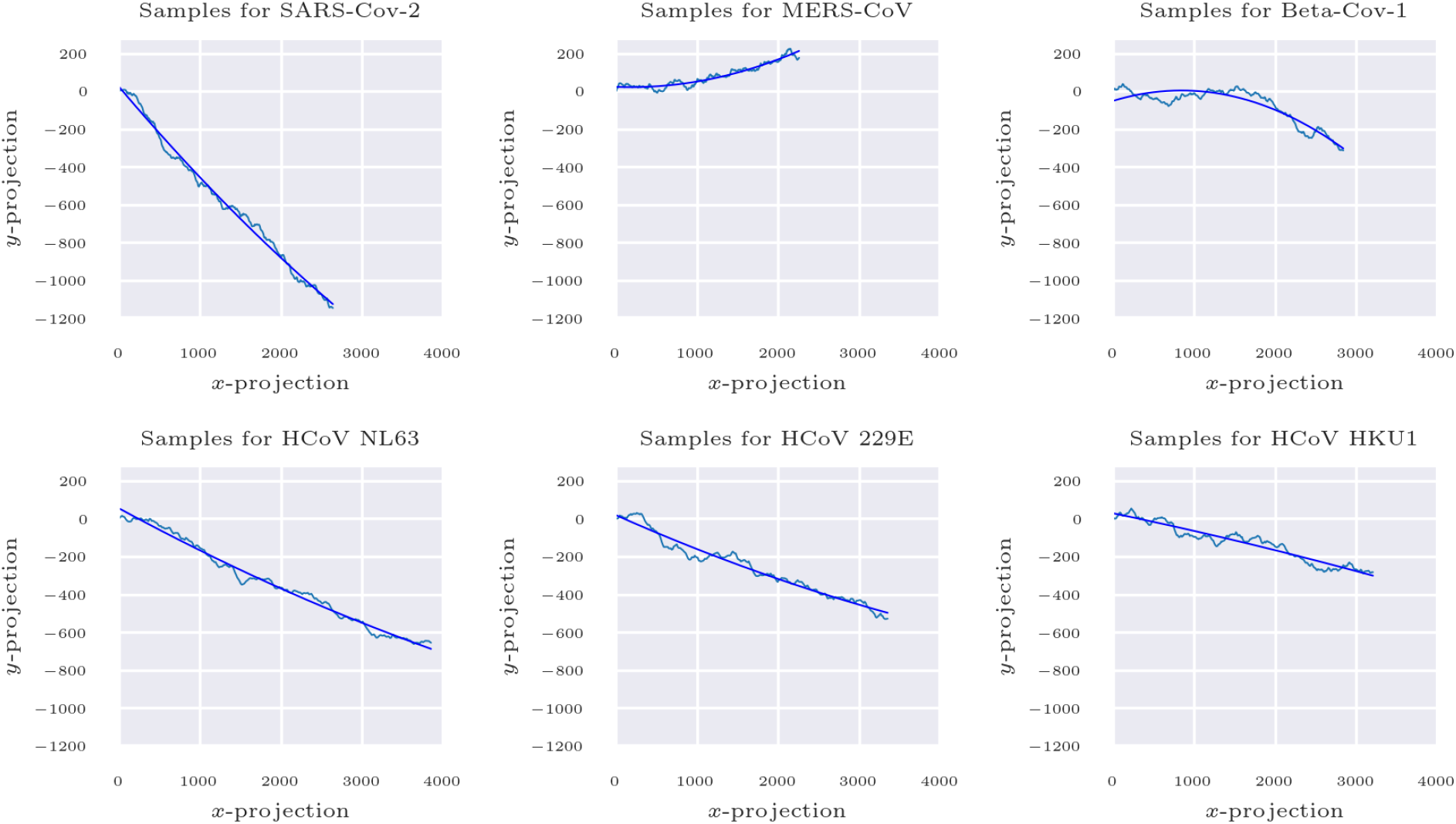
Depiction of a second-degree polynomial fit applied to samples representing each virus type, following a series of preprocessing steps, including Moving Average and uniform sampling.

Subsequently, PCA was applied for dimensional reduction, utilizing the computed matrices from both temporal and spectral domain features. The results are visually encapsulated and presented in Figure 7. The outcomes presented in Figure 7 are highly promising. A clear observation is that applying PCA to both temporal and spectral domain features facilitates robust discrimination among virus classes. Not only does it allow for effective differentiation, but it also reveals that the three most interrelated virus families appear close to each other within the PCA projection space, underscoring the method’s ability to maintain inherent relational proximities among the virus representations.

**Figure 7:**
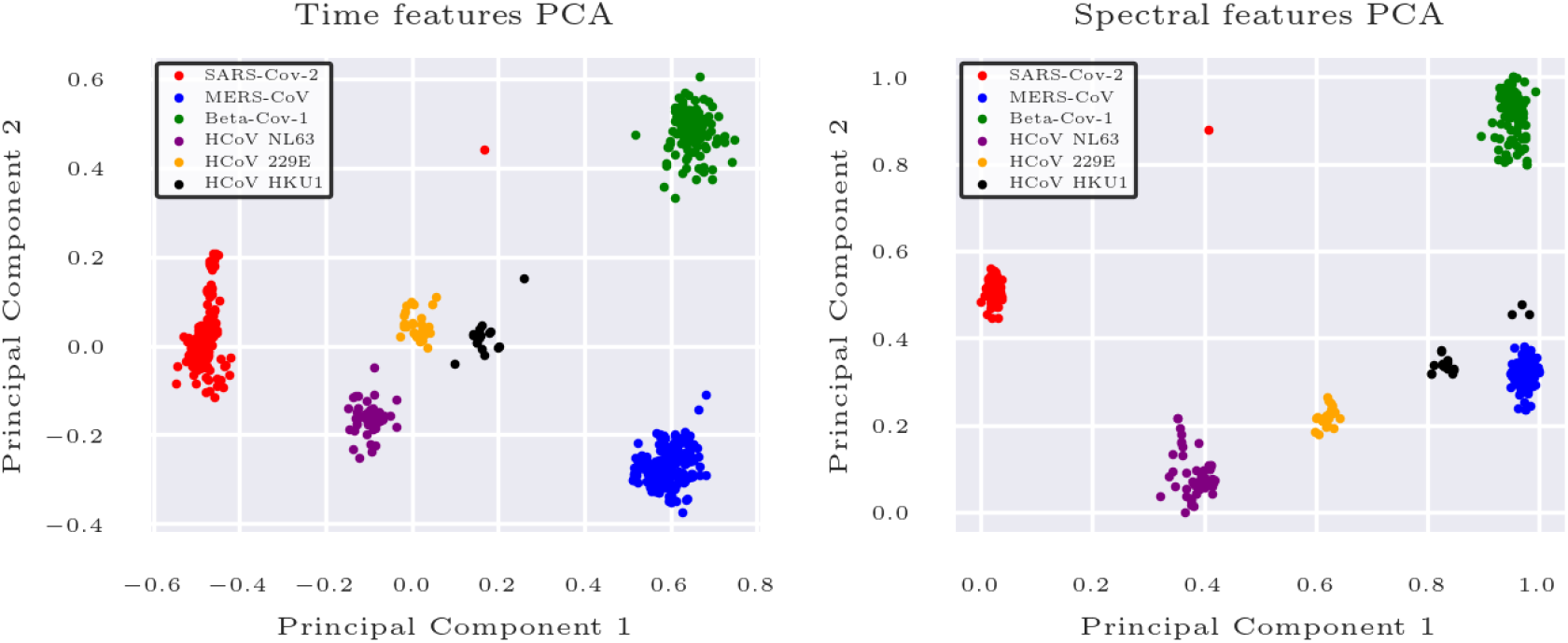
PCA Projection for Dimensionality Reduction. Panel A illustrates the application of PCA to the temporal domain features, demonstrating how data points are distributed in the principal component space. Panel B depicts the application of PCA to spectral domain features, showcasing the dispersion and grouping of data points in the reduced feature space.

### 3.1 Results of ML Models Used for Classification

Several performance metrics were employed to assess the efficacy of ML models in a multiclass classification challenge involving six distinct virus types from the Coronaviridae family—including SARS-CoV-2—. These include accuracy, precision, sensitivity, and F1-score, derived from both temporal and spectral data generated through the methodology introduced in this study. Utilizing *k*-fold cross-validation with *k* set to 10, we computed the mean of these metrics across each fold for the training dataset. The outcomes, showcasing the models’ performance based on spectral data, are detailed in Table 2.

**Table 2:** Performance metrics regarding test spectral data.

| Models | Accuracy | Recall | Precision | F1-score |
| --- | --- | --- | --- | --- |
| DT | 0.997 | 0.988 | 0.991 | 0.987 |
| RF | 0.997 | 0.996 | 0.993 | 0.993 |
| KNN | 0.998 | 0.999 | 0.998 | 0.999 |
| SVM | 1.000 | 1.000 | 1.000 | 1.000 |

The SVM demonstrated superior performance among the evaluated models, achieving optimal results across all metrics. This suggests its ability to differentiate all six viral subtypes in the training dataset. Following closely, the KNN model showcased near-optimal performance, with an accuracy of 99.99%, precision of 99.8%, recall of 99.9%, and an F1-score. The RF model also exhibited commendable performance, exceeding 99% across all metrics. Despite the DT model achieving accuracy and precision rates above 99%, it exhibited a marginal decline in recall (98.8%) and F1-score (98.7%) compared to the RF and KNN models. This indicates that the DT model might possess a slightly reduced capacity to identify all positive instances accurately. Thus, the KNN and SVM models display optimal and nearly optimal performance across all metrics. In contrast, the DT and RF models show minor reductions in recall, precision, and F1-score values for the training data.

Beyond the traditional performance metrics, standard deviation emerges as a vital statistic for assessing models’ variability, consistency, and stability throughout their training phase. For the RF, K-KNN, DT, and SVM models, the standard deviation values of accuracy were recorded as 0.00476, 0.00357, 0.00476, and 0.00, respectively. The SVM demonstrated a zero standard deviation, signaling uniform performance across all training folds, underpinning its exemplary metrics. Conversely, the KNN model presented the subsequent lowest standard deviation (0.00357), showcasing a high level of performance consistency during training. This low variability suggests that KNN’s performance metrics were notably stable across different training folds, contributing to the model’s overall reliability.

Regarding the RF and DT models, both achieved standard deviation values of 0.00476, reflecting comparable levels of consistency in their performance variability. This consistency suggests that these models and the previously mentioned KNN demonstrate stable prediction accuracy and are less prone to variations in the training data sets. Although the SVM recorded the lowest standard deviation (0.00), indicating unparalleled stability across training folds, this perfect uniformity may also hint at the model’s susceptibility to overfitting. To mitigate this risk and enhance the SVM’s ability to generalize across different data sets, a review and adjustment of its hyperparameters or an expansion of the training data might be warranted to diminish potential biases.

Figure 8 displays the confusion matrices for the test datasets of the models applied in this study, specifically for spectral data analysis. These matrices enable a direct visualization of the models’ generalization capabilities on data not encountered during the training phase. Importantly, this analysis encompasses all viral subtypes included in the database for the training phase, thereby illustrating the models’ ability to classify a broad range of viral genetic signatures accurately.

**Figure 8:**
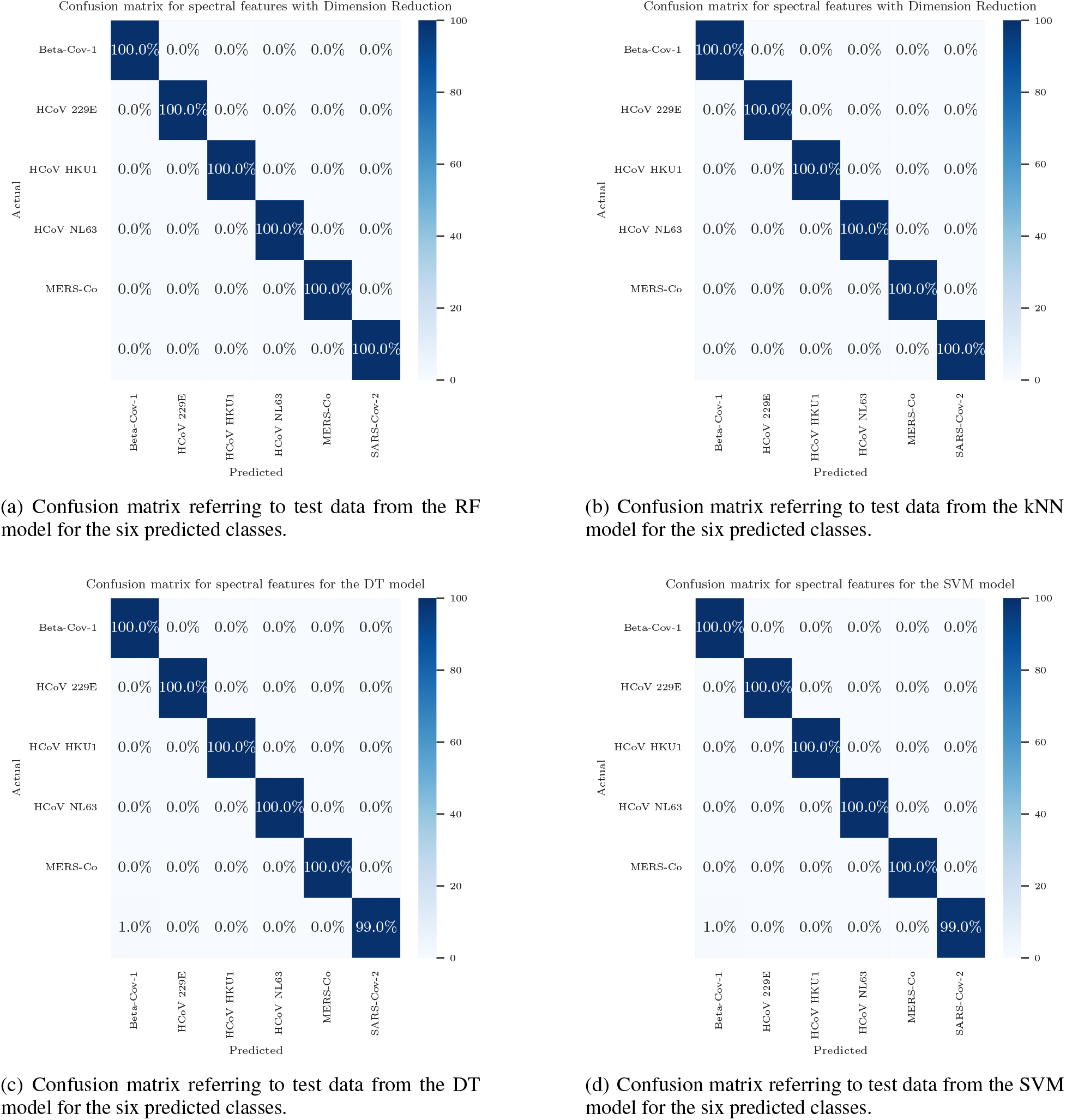
Confusion matrices for virus classification using spectral features. Each panel (a) through (d) corresponds to a different machine learning model—RF, kNN, DT, and SVM—and displays the predictive accuracy for six virus classes.

The proficiency of the RF and KNN models in accurately classifying all samples is prominently illustrated in the confusion matrices for the test data, as shown in Figure 8 a) and b)). This outcome underscores the efficacy of these models in differentiating between virus samples within the same viral family and correctly categorizing each sample into its respective class. The confusion matrix for the DT model, depicted in Figure 8 c), reveals a minor error where the model misclassified a single SARS-CoV-2 sample as Beta-CoV-1. This model achieved an accuracy of 99.50%, precision of 99.43%, recall of 99.83%, and an F1-score of 99.62% on the test data, aligning with its average training performance. Such a misclassification could stem from structural similarities between the virus classes, potentially due to shared genetic features that led to the model’s ambiguous interpretation. Figure 8 d) presents the confusion matrix for the SVM, which also faced challenges in accurately classifying a SARS-CoV-2 sample. On the test data, the SVM recorded slightly lower metrics compared to its training performance, with an accuracy of 99.50%, precision of 99.43%, recall of 99.83%, and an F1-score of 99.62%.

The same models previously utilized were applied to the temporal analysis data under the same training conditions as the previous phase. The performance metrics values for all models in this experiment can be viewed in Table 3.

**Table 3:** Performance metrics regarding test spectral data.

| **Models** | **Accuracy** | **Recall** | **Precision** | **F1-score** |
| --- | --- | --- | --- | --- |
| DT | 0.995 | 0.996 | 0.995 | 0.995 |
| RF | 0.996 | 0.995 | 0.990 | 0.991 |
| KNN | 0.998 | 0.999 | 0.998 | 0.999 |
| SVM | 0.998 | 0.991 | 0.994 | 0.991 |

Similar to the outcomes observed with spectral data analysis, all models demonstrated robust performance across all evaluated metrics, with accuracy, recall, precision, and F1-score ranging between 99.5% and 99.9%. However, the RF model exhibited slightly lower accuracy than the other models, yet it still managed to accurately identify a high percentage of actual positive instances. The standard deviation values associated with model training, reflecting accuracy, were 0.01430 for the DT, 0.00546 for RF, and 0.00357 for both KNN and SVM. KNN and SVM displayed identical standard deviation values, indicating consistent accuracy throughout their training. DT showed the highest standard deviation (0.01430), suggesting a more significant fluctuation in its accuracy than the other models.

The confusion matrices for the test dataset concerning temporal data are depicted in Figure 9. In contrast to the spectral data results, DT inaccurately classified samples from the MERS-CoV and SARS-CoV-2 classes, achieving accuracy, recall, precision, and F1-score of 99%, 96.09%, 99.51%, and 97.61%, respectively. Like the spectral analysis findings, RF and KNN accurately classified all samples, reaching peak performance metrics for the test data. Conversely, SVM misclassified a single sample (as shown in Figure 9 d)), achieving an accuracy of 99.43% on its test data.

**Figure 9:**
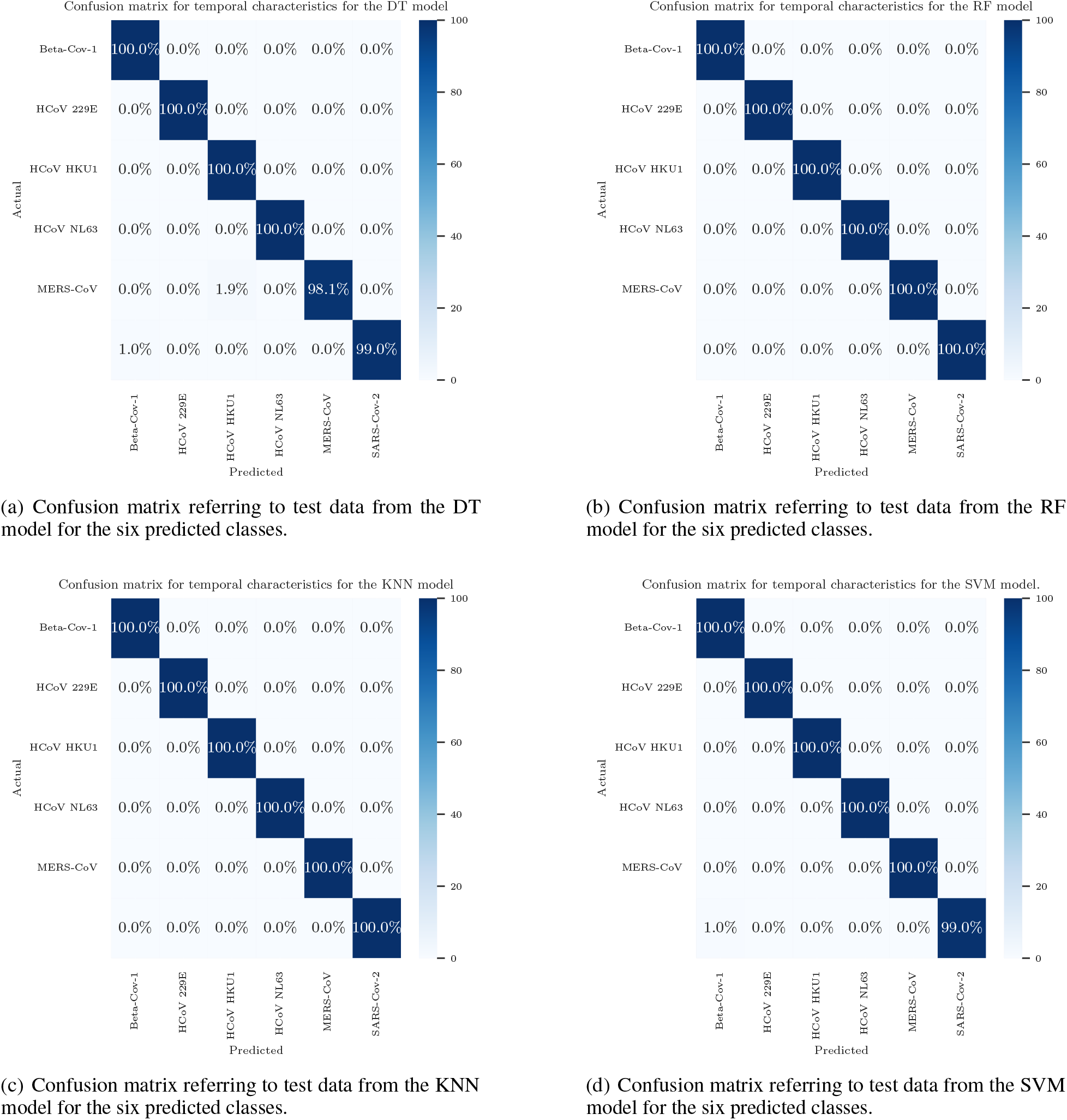
Confusion matrices for virus classification using temporal features. Each panel (a) through (d) corresponds to a different machine learning model—RF, kNN, DT, and SVM—and displays the predictive accuracy for six virus classes.

The evaluated models, DT, RF, KNN, and SVM, exhibited outstanding performance in a multiclass classification challenge involving six virus types from the Coronaviridae family, including SARS-CoV-2. With performance metrics exceeding 99%, these results underscore the proficiency of these models in differentiating among the various viral subtypes within the test datasets, as illustrated in Figure 7. This analytical representation enabled the identification of distinct characteristics among the evaluated classes despite their shared viral family lineage. It demonstrates that the employed genomic sequence processing technique effectively retains the unique attributes of each subgroup, affirming the method’s capability to accurately capture and represent the genetic diversity within the Coronaviridae family.

The remarkable proficiency of RF and KNN in accurately classifying all samples within the test data underscores their effectiveness, especially given the genetic similarities among the viral subtypes. While achieving accuracy and precision rates above 99%, the DT model experienced a minor reduction in recall and F1-score compared to RF and KNN. This indicates a potential limitation in its capacity to identify all positive cases, a capability that is vitally important in clinical scenarios where accurately distinguishing between various viral variants is critical for determining the correct treatment and intervention strategies. The spectral data analysis observed the highest training metrics and lowest standard deviation values, raising questions about the SVM’s ability to generalize. Across both spectral and temporal analyses, KNN and RF maintained low and consistent standard deviation values, aligning closely with the test data metrics. Interestingly, both SVM and DT misclassified a SARS-CoV-2 sample as belonging to the Beta-CoV-1 class. A closer examination of the spatial distribution of samples through principal component analysis (Figure 7) reveals a SARS-CoV-2 sample positioned near the Beta-CoV-1 class in both temporal and spectral domains. This proximity highlights the models’ nuanced understanding and utilization of characteristic data to make precise predictions despite the challenges presented by the genetic similarities among viruses.

Among studies utilizing ML models like K-KNN, SVM, RF, DT, and Extreme Gradient Boosting (XGBoost) for addressing SARS-CoV-2 viral classification challenges, accuracy rates spanned from 67% to 98%. This range underscores the adaptability and efficiency of these models in viral classification endeavors Dlamini et al. [2020], Singh et al. [2021b], Habib et al. [2020]. Specifically, in Dlamini et al. [2020], a perfect 100% performance across all metrics was observed for binary classification tasks among viruses of distinct species such as dengue, Ebola, tuberculosis, and some coronaviruses. This outcome was anticipated, given the significant phylogenetic differences between these groups. However, when extending the classification to a multiclass framework involving viruses from the same species but different continents, accuracy dropped to only 67.5%. This decline highlights the inherent challenges in differentiating between genetically similar entities.

While achieving outstanding performance metrics, the study utilized dinucleotide frequencies to characterize the virus, potentially limiting the model’s sensitivity in specific experiments. The research conducted by Singh et al. [2021b] employed 1,582 cDNA sequences from human, mammalian, and avian coronaviruses, alongside digital signal processing techniques, to undertake a binary classification task distinguishing between SARS-CoV-2 and non-SARS-CoV-2 samples. This analysis involved 615 SARS-CoV-2 and 967 non-SARS-CoV-2 samples, applying KNN, SVM, DT, and RF models. The Random Forest model exhibited the highest precision, achieving 98% for the training set and 97.47% for the test set, indicating superior performance. In contrast, the SVM showed reduced capacity for generalization. Notably, despite RF recording the third lowest performance in our investigation, the SVM demonstrated accuracy exceeding 99% for training and test datasets, showcasing its practical application in viral classification tasks.

In their study, Habib et al. [2020] tackled a multiclass classification challenge involving the same viral subtypes discussed in this paper, leveraging genomic sequences for analysis. They utilized a DL approach, specifically a Multi-layer Perceptron classifier, to achieve precision and recall metrics ranging from 1 to 0.99. While potent in its predictive capabilities, this DL model incurs a significantly higher computational cost than traditional ML techniques. A total of 1925 genome sequences were analyzed, with data preprocessing employing a natural language processing (NLP) method. This method converts textual information into a vector, relying on the frequency (count) of each word appearing in the text. While effective in many contexts, this approach may pose challenges in accurately capturing viral mutations, as the evolutionary dynamics of viruses can alter their genomic sequences.

In their investigation, Murugaiah and Ganesan [2022] employed a signal processing technique for preprocessing to classify seven coronavirus strains, analyzing 1000 data samples across six different classifiers. The models evaluated included the Convolutional Neural Network (CNN), Artificioal Neural Network (ANN), KNN, and SVM, with the latter two yielding the most impressive accuracy rates of 97.96%, 93.60%, 92.80%, and 91.84%, respectively. A separate study by Randhawa et al. [2020] utilized six ML algorithms to classify entities at various taxonomic levels, achieving an outstanding 98.1% accuracy with the SVM model for classifying between four genera, based on a dataset comprising only 208 sequences from the coronaviridae family. However, the relatively small dataset size may pose challenges in terms of model generalization and performance across broader applications.

Numerous studies on multiclass classification within the coronaviridae family’s viral subtypes have leveraged the same database as this current investigation, showcasing results surpassing 90% accuracy. These studies predominantly utilized DL methodologies or concentrated on determining the presence or absence of SARS-CoV-2 Singh et al. [2021b], Tampuu et al. [2019], Murugaiah and Ganesan [2022], Dlamini et al. [2020]. Among these, the technique proposed in this paper achieved superior and more consistent outcomes, with Habib et al. [2020] being a notable exception, where a 100% accuracy was reached for the same classification challenge using DL approaches. Despite their effectiveness, DL techniques are known for their significant computational demands, surpassing those of more straightforward ML methods like KNN, SVM, and RF. Furthermore, DL models often necessitate larger datasets for training or more detailed representations of the viral genomes to perform optimally.

Overall, the selection of preprocessing techniques such as frequency-based methods, GSP, NLP, and the approach proposed in this study (Dlamini et al. [2020], Habib et al. [2020], Murugaiah and Ganesan [2022]) might not fully capture the intricacies involved in classifying viruses within the same species. For a more equitable assessment, additional factors should be considered, including the preprocessing time of sequences and models and the often omitted details of model hyperparameters, which are crucial for enabling the replication and testing of methodologies. Enhancing the model’s generalization capability necessitates an expansion of the dataset size, particularly for underrepresented virus subtypes. Nonetheless, open genomic databases face a notable constraint: they predominantly feature sequences related to SARS-CoV-2 due to the pandemic, resulting in a scarcity of sequences for other coronaviridae family subtypes. This significant limitation must be considered when interpreting the findings and applying these methodologies to broader and more complex classification challenges, such as distinguishing viruses across different taxonomic levels.

## 4. Conclusion

The complexity and variability inherent in genomic data introduce significant challenges in extracting and interpreting features, not to mention the considerable computational demands required for processing. The way genomic data is pre-processed and transformed prior to application in standard machine learning techniques directly impacts the outcomes of classification efforts. Thus, the quality of genomic data representation is crucial for generating precise and meaningful insights from these models. This study presents a novel approach to encoding cDNA sequences into unit vector transitions in a two-dimensional space. Our primary objective is to illuminate the potential of this method to enhance the application of conventional machine learning models in the classification of six virus types from the same viral family. By requiring minimal hardware resources and achieving performance metrics between 96% and 99%, the proposed encoding method is a computationally efficient and promising enhancement to current DNA sequence encoding techniques. Its advantages include simplicity, visual interpretability, and ease of implementation, making it a potentially invaluable complement to existing strategies in the field.

The significance of the proposed method in advancing virus classification and gene sequence analysis cannot be overstated. It offers a robust tool for scientists and researchers, simplifying the complexities of genomic data and improving the efficacy and efficiency of machine learning models in genomic studies. This innovative data representation method marks a notable leap forward in our ability to classify viruses accurately, showcasing high accuracy in distinguishing between different types within the coronaviridae family. Furthermore, by substantially reducing the computational resources needed for such analyses, this method makes advanced genomic analysis accessible, even in environments constrained by limited resources.

The practical implications and potential benefits of adopting this novel approach are far-reaching, especially in real-world scenarios where rapid and accurate virus detection is crucial for effective disease management and outbreak control. The method’s simplicity and computational efficiency make it particularly suitable for quick viral classification, aiding in timely decision-making during public health emergencies. By bridging the gap between complex genomic data and the applicability of machine learning, this study contributes significantly to bioinformatics and public health, offering a vital resource in the ongoing efforts to understand and combat viral diseases. The comparison with other GSP techniques further validates the effectiveness of the proposed method, underscoring its importance as a cutting-edge tool in the scientific community’s arsenal against viral outbreaks.

## Data Availability

All data produced in the present study are available upon reasonable request to the authors

## Notes

### Competing Interest Statement

The authors have declared no competing interest.

### Funding Statement

This study did not receive any funding

### Author Declarations

The study uses DNA sequences extracted from the 2019 Novel Coronavirus Resource (2019nCoVR), which is an open repository accessible at https://bigd.big.ac.cn/ncov/.

### Summary of Updates

This version of the manuscript has undergone revision to enhance certain elements. Specifically, figure captions have been amended to rectify initial incompleteness, additional references have been incorporated to bolster the foundational literature of the study, and minor textual revisions have been implemented throughout the manuscript.

